# Pharmacogenetic Characterization of Cytochrome P450 Genes involved in Psychotropic Medication Metabolism in a Cohort of Patients with Prader-Willi Syndrome

**DOI:** 10.64898/2026.05.09.26352521

**Authors:** Alex Moreno-Armengol, Rocío Pareja, Alba Hernández-Lázaro, Laura Capel, Raquel Corripio, Assumpta Caixàs, Neus Baena

## Abstract

Prader-Willi syndrome (PWS) is a rare multisystemic disorder characterized by obesity, endocrine dysfunctions, and psychiatric comorbidities, which imply frequent use of psychotropic medications. They account for atypical responses to standard dosages of psychiatric drugs. Pharmacogenetics could be part of the reason for this situation, potentially offering a valuable tool for individualized treatment.

This study analyzed allelic and phenotypic frequency distributions of five of the main cytochrome P450 enzymes (CYP2D6, CYP2B6, CYP2C19, CYP2C9, CYP3A4) involved in psychiatric drug metabolism in 47 patients with genetically confirmed diagnosis of PWS and compared them to reference frequencies in the general European population.

Allelic frequency comparisons between the European reference population and the overall PWS cohort revealed a significant global difference for CYP2B6, with CYP2C19 and CYP2D6 showing trends toward significance. Although no global allelic differences remained significant after false discovery rate correction, post-hoc analyses consistently identified an enrichment of reduced- or non-functional alleles CYP2B619 and CYP2D610 in patients with PWS. Predicted metabolizer phenotype analyses showed a significant shift toward intermediate metabolizers of CYP3A4 in the PWS cohort, with corresponding depletion of normal metabolizers. Subgroup analyses indicated that allelic differences were more pronounced in maternal uniparental disomy and non-deletion subtypes, particularly for CYP2B6, although no significant differences were observed between PWS genetic subtypes.

Overall, results imply potential differences in metabolizing activity in PWS patients, and subsequent implications in drug efficacy and tolerability. These results support the idea that pharmacogenetic testing may improve therapeutic decision-making in PWS for psychiatric treatment. Larger studies are needed to confirm these preliminary results.

## 1. Introduction

Prader-Willi syndrome (PWS) is a rare multisystemic disorder with an estimated prevalence of one in 10.000 to 30.000 individuals (Cassidy et al., 2012). It is recognized as the most common form of syndromic obesity. PWS is caused by the loss of expression of paternally inherited genes in the imprinted region of chromosome 15q11.2–q13. This loss is primarily caused by paternal deletions (DEL, 65–75%), maternal uniparental disomy (UPD, 20–30%) or imprinting defects (ID, 5%) (Butler et al., 2018).

The main clinical characteristic of PWS is hyperphagia, which is caused mainly by lack of satiety. It can lead to progressive weight gain and finally obesity that constitutes the main cause of morbidity and mortality in PWS. Other common manifestations include severe neonatal hypotonia, feeding difficulties and low weight gain during early infancy, mild cognitive impairment, motor and language developmental delays, hypothalamic dysfunction that leads to endocrine dysregulations (hypogonadism, growth hormone deficiency, etc.), sleep abnormalities, scoliosis, among others (Cassidy et al., 2012; Butler et al., 2019).

Behavioral and psychiatric symptoms are another core feature of the syndrome. Patients exhibit a characteristic behavioral profile with anxiety, impulsiveness, temper tantrums, obsession, cognitive rigidity, psychosis and autolesions (mainly skin picking), among others, since early childhood. These behavioral dysregulations are exacerbated during adolescence, when also psychiatric conditions like psychosis and mood disorders may need psychotropic drugs (if not initiated previously) and require sustained treatment through adulthood (Guinovart et al., 2019; Schwartz et al., 2021; Aman et al., 2024).

Despite psychotropic drugs have shown limited efficacy in achieving behavioral control of the patients, their use is very frequent, typically combining different drugs simultaneously (Bonnot et al., 2016; Butler et al., 2019). In addition, patients may receive concomitant pharmacological treatment to manage other comorbidities (diabetes, dyslipidemia, hypothyroidism…) (Butler et al., 2019), that may increase the risk of drug-drug interactions and side-effects.

Pharmacogenetics offers a promising approach to guide treatment decisions in the context of personalized medicine. This discipline examines how genetic variations on genes coding for drug-metabolizing enzymes affect pharmacokinetics and treatment response. Among all this group of enzymes, cytochrome P450 enzyme family (CYP), a heme-based superfamily of proteins that perform oxidative phosphorylation of drugs and toxins among other functions, emerges as one of the most relevant members (Samer et al., 2013). The CYP enzymes, located primarily in the liver, are responsible for metabolizing over 80% of all drugs through oxidative reactions (Wilkinson, 2005; Zanger and Schwab, 2013). Drugs that undergo metabolism may act as substrate for more than one CYP enzyme, and some may require a first-pass metabolism to become pharmacologically active. Also, some drugs can act as inhibitors or inducers of some CYP enzymes.

Psychotropic drugs are mainly metabolized by CYP2D6, CYP2B6, CYP3A4, CYP2C9, CYP2C19 and CYP1A2 enzymes (Gross and Daniel, 2018). Several psychotropic medications have gene-drug interactions described in the main pharmacogenetic repositories, mainly involving enzymes CYP2D6 and CYP2C19. Guidelines included provide genotype-based recommendations for drug selection and dosage. Despite their scientific validation and international adoption, Spanish National Health System still does not integrate these guidelines. The Spanish national portfolio of Genetic Studies Services has already created the Pharmacogenomics studies, but implementation remains pending. Expanding evidence in underrepresented populations, like in PWS, may give a proof of concept to start using genotyped-based psychiatric pharmacological selection and dosage and move towards personalized medicine.

Genetic polymorphisms in CYP450 encoding genes may alter enzymatic activity. Depending on the effect, individuals can be classified into poor metabolizers (PM; little/no function), intermediate metabolizers (IM; decreased function), normal metabolizers (NM; normal activity), or rapid/ultra-rapid metabolizers (RM/UM; increased activity) (Caudle et al., 2017; Tayeh et al., 2022). These phenotypes can influence both drug efficacy and the risk of adverse events at standard dosages. A prior study reported differences in phenotypic distributions of these genes in northamerican individuals with PWS compared to the general population, suggesting them as a potential reason for atypical psychotropic treatment outcomes and difficulties for dose-adjusting in PWS (Forster et al., 2021).

The aim of this research is to determine allelic and phenotypic frequency distributions of CYP2D6, CYP2B6, CYP3A4, CYP2C9, CYP2C19 genes in a cohort of 47 Spanish patients with PWS, and compare them to general European population data, to identify differences that may justify the observed atypical responses to psychiatric drugs.

CYP1A2 gene analysis has not been included, despite being responsible for the processing of psychiatric drugs like haloperidol, melatonin or olanzapine, due to scarce data on European population allelic and phenotypic frequencies. A comprehensive list of analyzed alleles and their defining polymorphisms is provided in Table 1.

**Table 1.** Comprehensive list of all alleles and polymorphisms included.

**CYP2B6**
| **Allele** | **Polymorphism(s) (rs code)<sup>1</sup>** |
| --- | --- |
| *1 | None (Wild-Type) |
| *2 | rs8192709 |
| *3 | rs45482602 |
| *4 | rs2279343 |
| *5 | rs3211371 |
| *6 | rs3745274; rs2279343 |
| *7 | rs3745274; rs2279343; rs3211371 |
| *8 | rs12721655 |
| *9 | rs3745274 |
| *18 | rs28399499 |
| *19 | rs3745274; rs2279343; rs34826503 |
| *22 | rs34223104 |

**CYP2C9**
| **Allele** | **Polymorphism(s) (rs code)<sup>1</sup>** |
| --- | --- |
| *1 | None (Wild-Type) |
| *2 | rs1799853 |
| *3 | rs1057910 |
| *5 | rs28371686 |
| *6 | rs9332131 |
| *8 | rs7900194 |
| *9 | rs2256871 |
| *11 | rs28371685 |
| *12 | rs9332239 |
| *13 | rs72558187 |
| *15 | rs72558190 |

**CYP2C19**
| **Allele** | **Polymorphism(s) (rs code)<sup>1</sup>** |
| --- | --- |
| *1 | *rs3758581* |
| *2 | *rs3758581*; rs4244285; rs12769205 |
| *3 | *rs3758581*; rs4986893 |
| *4 | *rs3758581*; rs28399504 |
| *5 | *rs3758581*; rs56337013 |
| *8 | *rs3758581*; rs41291556 |
| *9 | *rs3758581*; rs17884712 |
| *10 | *rs3758581*; rs6413438 |
| *12 | *rs3758581*; rs55640102 |
| *14 | *rs3758581*; rs55752064 |
| *17 | *rs3758581*; rs12248560 |
| *22 | *rs3758581*; rs140278421 |
| *24 | *rs3758581*; rs118203757 |

**CYP2D6**
| **Allele** | **Polymorphism(s) (rs code)<sup>1</sup>** |
| --- | --- |
| *1 | None (Wild-Type) |
| *2 | rs16947; rs1135840; rs1080985<sup>2</sup> |
| *3 | rs35742686 |
| *4 | rs3892097 |
| *5 | Full Gene Deletion |
| *6 | rs5030655 |
| *9 | rs5030656 |
| *10 | rs1065852; rs1135840 |
| *17 | rs28371706; rs16947; rs1135840 |
| *29 | rs59421388; rs61736512 + *rs1058164*;<br>rs16947; rs1135840 |
| *34 | rs16947 |
| *39 | rs1135840 |
| *41 | rs28371725; rs16947; rs1135840 |

CYP3A4
| Allele | Polymorphism(s) (rs code)<sup>1</sup> |
| --- | --- |
| *1 | None (Wild-Type) |
| *1B | rs2740574 |
| *6 | rs4646438 |
| *7 | rs56324128 |
| *18 | rs28371759 |
| *20 | rs67666821 |
| *22 | rs35599367 |
| *26 | rs138105638 |
| *30 | rs778013004 |

<sup>1</sup> Data obtained from www.pharmvar.org/genes/.
<sup>2</sup> rs1080985 corresponds to allele \*2A, a variant of allele \*2.
Polymorphisms rs3758581 (*CYP2C19*) and rs1058164 (*CYP2D6*), in italics, were not included into the OpenArray panel due to failure of the probes when designing it.

## 2. Materials and Methods

### 2.1. Patients

A total of 47 patients with PWS were recuited from the Endocrinology Department of a reference center (Corporació Sanitària Parc Taulí, Sabadell, Barcelona, Spain). Patients underwent a complete medical and clinical assessment. Inclusion criterion was a genetically confirmed diagnosis of PWS with MS-MLPA technique (MRC Holland). Sex and age were not considered in recruitment, since we are studying genetic polymorphisms in autosomes. The cohort was composed of Spanish individuals, almost exclusively of Caucasian ethnicity.

The study protocol was approved by the local ethics committee (Institutional Committee of Ethics and Investigation with Drugs Parc Taulí in Sabadell, Barcelona; ref. 2024/5109). The study complied with the Declaration of Helsinki, Good Clinical Practice (ICH-GCP). All patients agreed to participate after being informed together with their parents or caregivers and signed an assent or a consent; their parents/legal guardians signed the consent form before enrolment if appropriate.

### 2.2 DNA extraction

A blood sample from each patient was obtained during rutinary control visits. Blood was placed in an EDTA-anticoagulating tube. Genomic DNA was then extracted using the Maxwell® CSC AS6000 (PROMEGA) Device and the Maxwell® RSC Blood DNA Kit (Promega) both of them CE-IVD certified, following the manufacturer’s instructions. DNA was eluted in 50–100 µL of elution buffer and stored at −20 °C until analysis.

### 2.3. DNA quality assessment

DNA concentration and purity were assessed using a NanoDrop™ spectrophotometer (Thermo Fisher Scientific), measuring concentration of DNA in ng/µL and the quality parameters using absorbance at 260/280 nm and 260/230 nm.

### 2.4. Custom polymorphism panel design

Genotyping was performed using TaqMan® OpenArray® Genotyping Plates (Thermo Fisher Scientific). Panel format was 60 single-nucleotide polymorphisms (SNP) per sample. The panel was custom designed. Target alleles were decided based on both recommendations from the ClinPGx repository (www.clinpgx.org), and the alleles analyzed in Forster et al. previous work. A list of alleles included can be seen in Table 1.

### 2.5. Polymorphism determination

TaqMan® OpenArray® Genotyping Plate (Thermo Fisher Scientific) was genotyped following the manufacturer’s protocol. DNA concentration in samples used was 50 ng/µL. Samples were loaded onto arrays using the AccuFill™ System, and amplification and allele detection were carried out on a QuantStudio™ 12K Flex Real-Time PCR System. Genotypes were called using ThermoFisher® Cloud Software.

### 2.6. Copy Number Variations Assessment

CNV assessment was performed using MLPA P128 CYP450 kit (MRC Holland) following manufacturer’s protocol. Fragment analysis for peak quantification was then performed with SeqStudio (Thermo Fisher Scientific). The fragment analyses were interpreted using GeneMapper™ software 6 (Thermo Fisher Scientific, Waltham, MA, USA) and the analysis of the results was performed using a customized Excel spreadsheet.

### 2.7. Genotyping results analysis

Genotyping results from each sample were then analyzed to determine the alleles present in each individual. Genotype to phenotype association was done using ClinPGxreference tables for CYP2B6, CYP2C9, CYP2C19, CYP2D6, available at https://www.clinpgx.org/page/pgxGeneRef. For CYP3A4, this association was done through inference of each allele predicted metabolic activity, obtained from reference bibliography.

### 2.8. Statistical analysis

Frequencies for general population were obtained also from ClinPGx repositories (https://www.clinpgx.org/page/pgxGeneRef) for CYP2B6, CYP2C9, CYP2C19, CYP2D6. CYP3A4 lacked reference tables, thus, allelic frequencies were obtained from the 1000 Genomes repository (https://www.internationalgenome.org/), and Zhou et al. work was used to obtain phenotypic frequencies (Zhou and Lauschke, 2022). Allele *2A of CYP2D6 was considered equivalent to *2 in this study, in accordance with ClinPGx conventions. The sample size of all groups can be seen in Table 2.

**Table 2.** Sample sizes of European and PWS cohorts, and PWS subtypes. For European populations, data was obtained from ClinPGx reference tables (https://www.clinpgx.org/page/pgxGeneRef) for all genes except CYP3A4, whose data for alleles (1) and phenotypes (2) were obtained from different sources.

| Gene | European reference (n) | PWS total (n) | DEL (n) | UPD (n) | Non-DEL (n) |
| --- | --- | --- | --- | --- | --- |
| CYP2B6 | 69572 | 45 | 27 | 15 | 18 |
| CYP2C19 | 71782 | 45 | 27 | 15 | 18 |
| CYP2D6 | 29114 | 45 | 27 | 15 | 18 |
| CYP2C9 | 65090 | 47 | 28 | 15 | 19 |
| CYP3A4 | 1006<sup>1</sup><br>77165<sup>2</sup> | 46 | 27 | 15 | 19 |

Allelic and predicted metabolizer phenotype distributions for each gene were compared between individuals with Prader–Willi syndrome (PWS) and the European reference population. The PWS cohort was further stratified according to genetic subtype, and comparisons with the European population were performed separately for individuals with DEL, UPD, and a non-deletion group (Non-DEL), which comprised UPD cases and patients with other non-deletion molecular mechanisms, including ID. In addition, direct comparisons were conducted between DEL and UPD subgroups, as well as between DEL and Non-DEL subgroups.

For allelic frequency analyses, only alleles included in the genotyping panel that were either observed in the PWS cohort and/or had a reported prevalence >1% in the reference population were included in the statistical analyses. Alleles not meeting these criteria were excluded from testing. Predicted metabolizer phenotypes were classified as ultrarapid (UM), rapid (RM), normal (NM), intermediate (IM), or poor (PM) metabolizers. Depending on the gene analyzed, only a subset of these phenotype categories was observed or biologically defined, and analyses were therefore restricted to the phenotypes applicable to each gene.

For each gene, global comparisons of allelic and phenotype frequency distributions between population pairs were conducted using contingency tables (r×2). Since in all global comparisons, at least 1 expected cell count was smaller than 5, Fisher’s exact test with Monte Carlo simulation (10[ iterations) was applied to obtain stable p-value estimates for high-dimensional tables. [34] False discovery rate (FDR) correction for multiple testing was applied separately for allelic and phenotype analyses using the Benjamini–Hochberg procedure across genes within each comparison. Adjusted p-values (q-values) <0.05 were considered statistically significant.

Post-hoc analyses were performed in all genes, despite not showing global differences, due to the exploratory intention of the present study. Exploratory post-hoc analyses were performed to identify categories potentially driving distributional shifts. Each allele or phenotype category was tested against all remaining categories combined using 2×2 Fisher’s exact tests (exact method).

Within-gene multiple testing in post-hoc analyses was controlled using the Benjamini–Hochberg False Discovery Rate (FDR) procedure. Post-hoc results were interpreted as exploratory and were contextualized with respect to the corresponding global test results.

Statistical significance was determined by p-value smaller than 0,05 in all tests. All statistics and graphical representations were done using R Statistics (version 4.4.1).

## 3. Results

The results obtained according to the different categories are described below.

### 3.1. European reference population vs PWS patients

#### 3.1.1. Allele frequency distributions

Global comparisons of allele frequency distributions between the European reference population and the overall PWS cohort revealed statistically significant differences for CYP2B6 (p < 0.01) and CYP2D6 (p = 0.02), while CYP2C19 (p = 0.06) showed a trend toward significance. No statistically significant global differences were detected for CYP2C9 and CYP3A4. No allelic distribution comparison remained significant after FDR correction (Figures 1A-5A; Supplementary Table 1).

**Figure 1.**
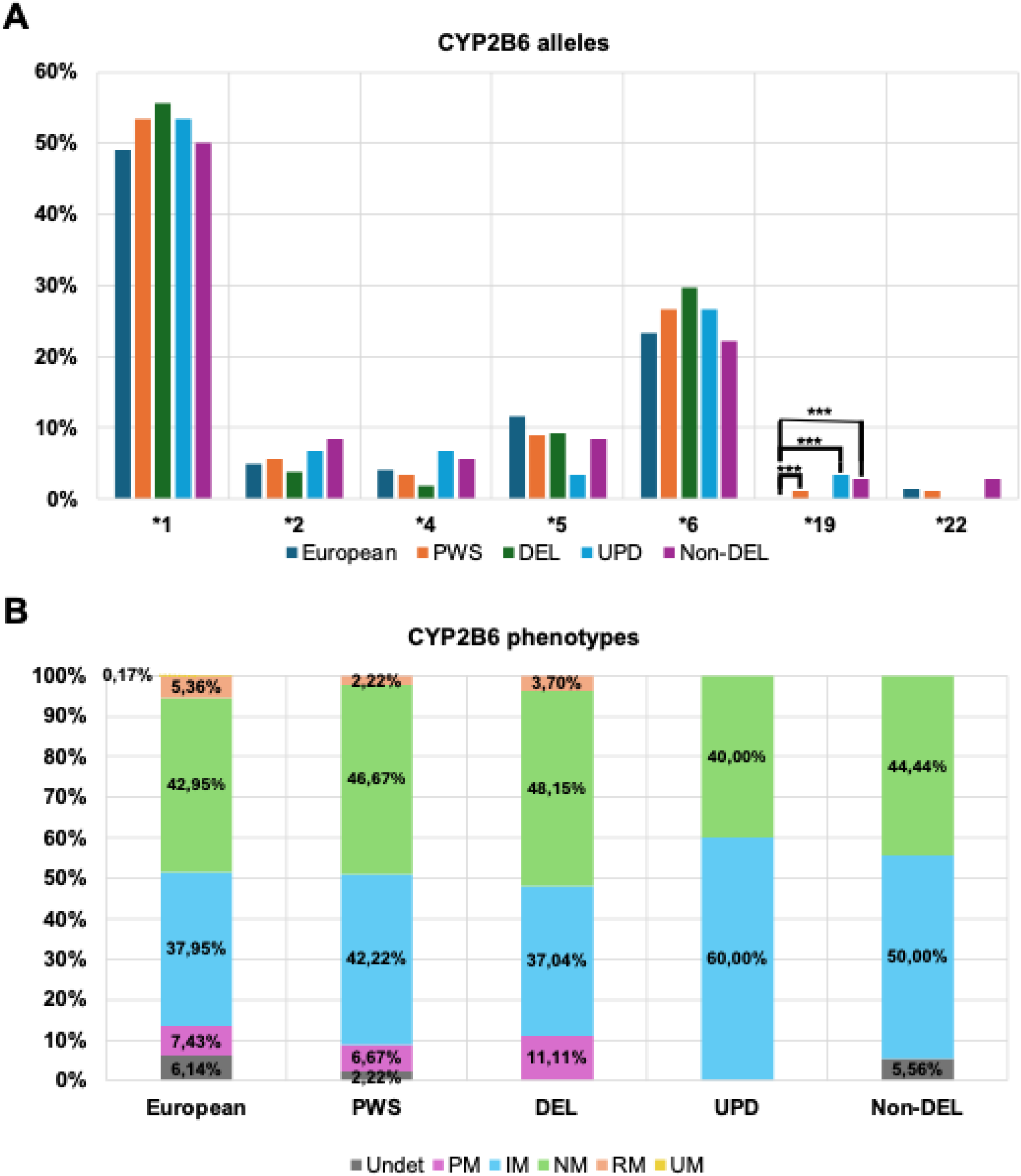
CYP2B6 allelic and predicted metabolizer phenotype distributions across study groups. (A) Allele frequency (%) in the European reference population and in the PWS cohort, including genetic subtypes. Global distribution comparisons showed significant differences in European vs. PWS (p < 0.01), European vs. UPD (p < 0.001) and European vs. Non-DEL (p < 0.01). Significant post-hoc differences are indicated as following: *** (p < 0.001). (B) Distribution of predicted metabolizer phenotypes. No global comparisons, nor post-hoc comparisons showed significant differences.

**Figure 2.**
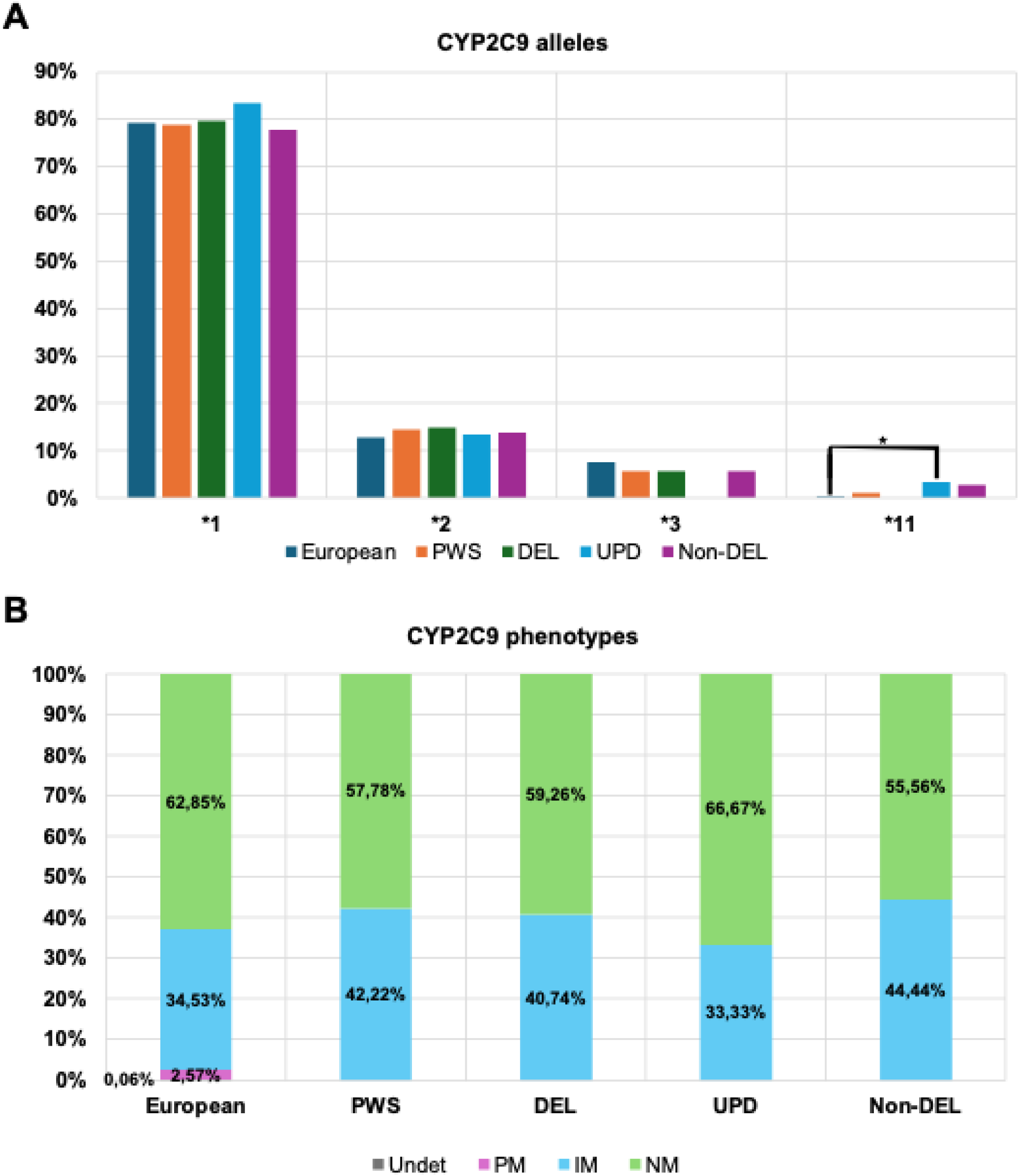
CYP2C9 allelic and predicted metabolizer phenotype distributions across study groups. (A) Allele frequency (%) in the European reference population and in the PWS cohort, including genetic subtypes. Global distribution comparisons showed significant differences in European vs. UPD (p < 0.05). Significant post-hoc differences are indicated as following: * (p < 0.05). (B) Distribution of predicted metabolizer phenotypes. No global comparisons, nor post-hoc comparisons showed significant differences.

**Figure 3.**
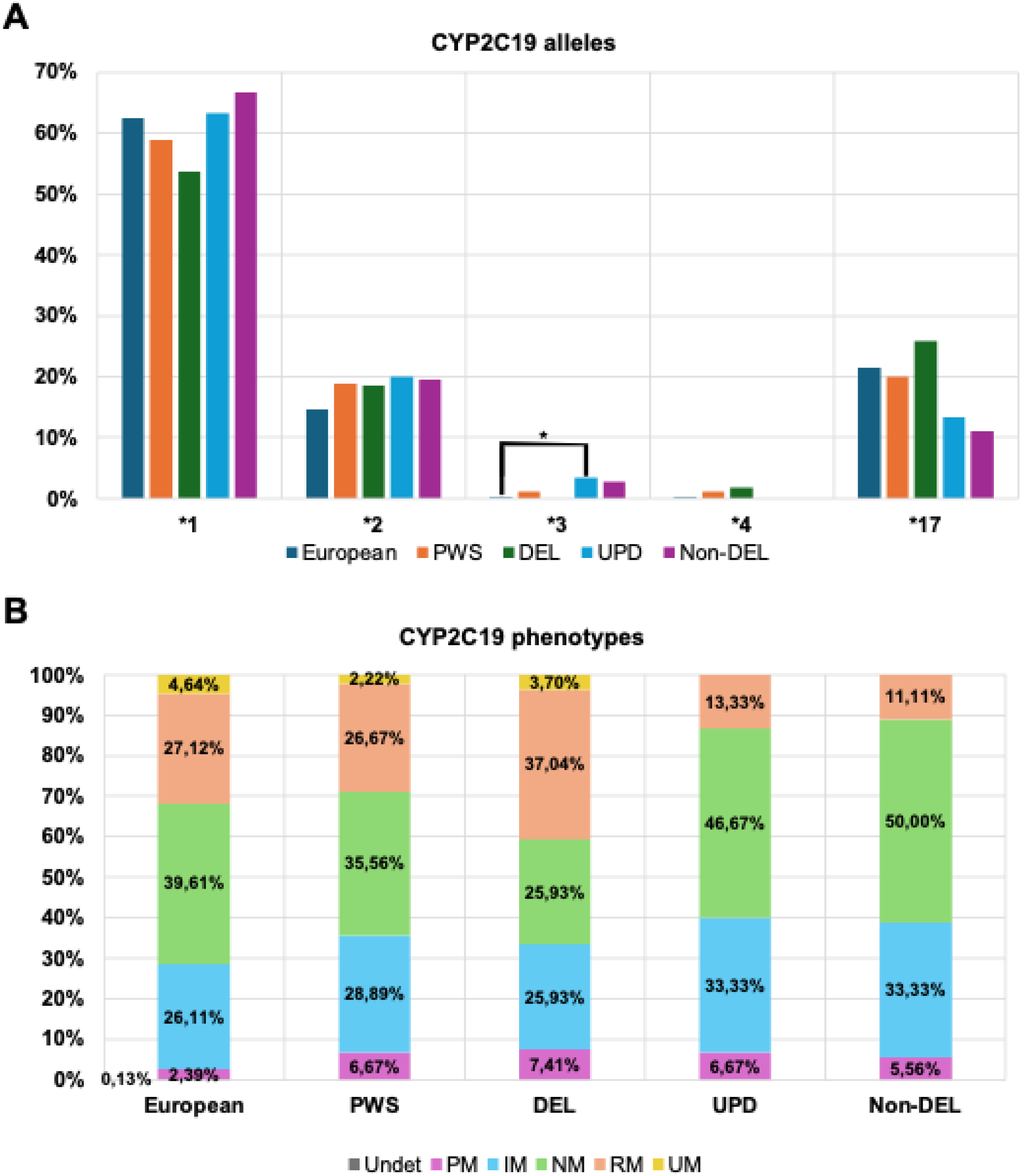
CYP2C19 allelic and predicted metabolizer phenotype distributions across study groups. (A) Allele frequency (%) in the European reference population and in the PWS cohort, including genetic subtypes. Global distribution comparisons showed significant differences in European vs. Non-DEL (p < 0.05). Significant post-hoc differences are indicated as following: * (p < 0.05). (B) Distribution of predicted metabolizer phenotypes. No global comparisons, nor post-hoc comparisons showed significant differences.

**Figure 4.**
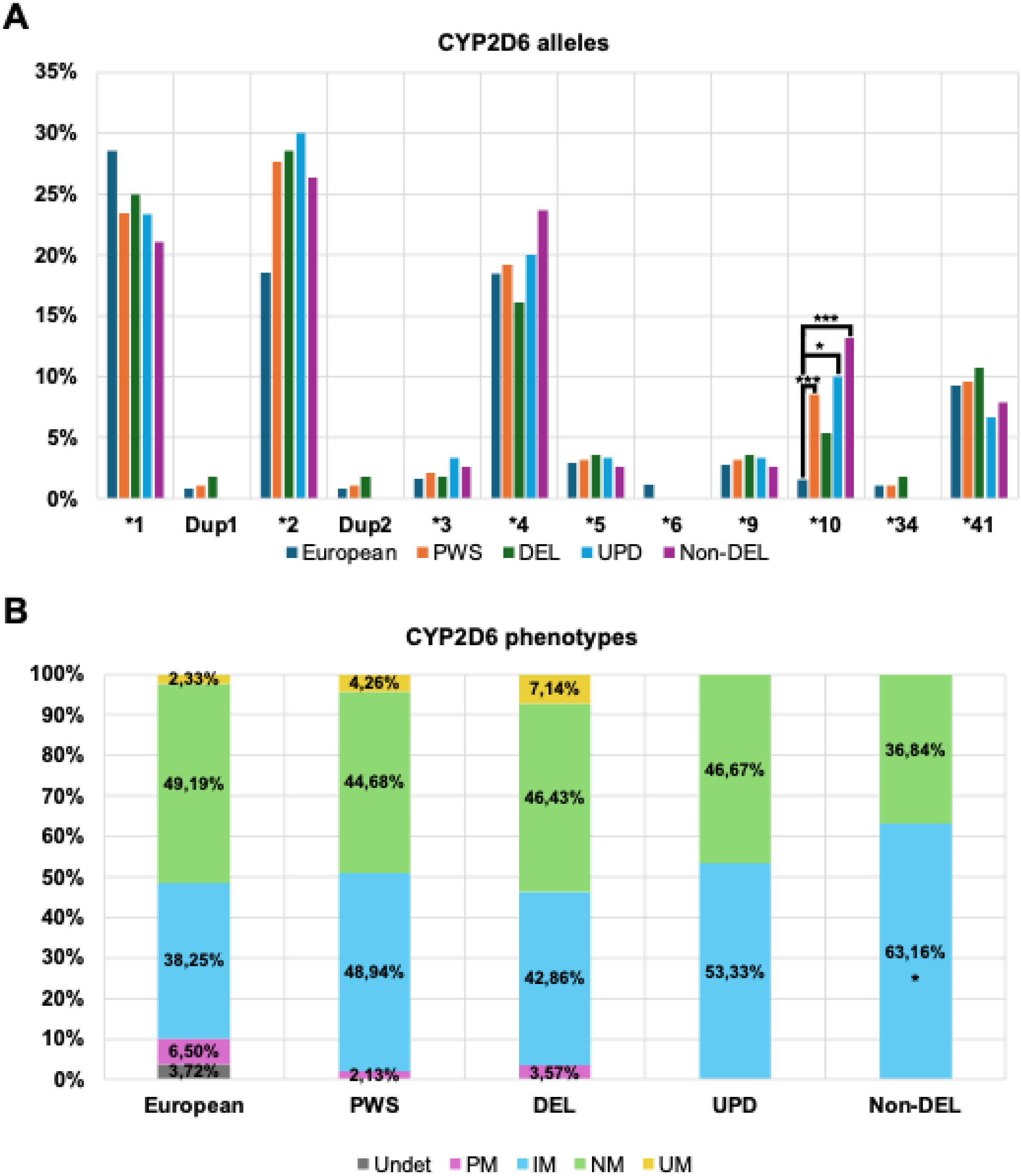
CYP2D6 allelic and predicted metabolizer phenotype distributions across study groups. (A) Allele frequency (%) in the European reference population and in the PWS cohort, including genetic subtypes. Global distribution comparisons showed significant differences in European vs. PWS (p < 0.05). Significant post-hoc differences are indicated as following: * (p < 0.05); *** (p < 0.001). (B) Distribution of predicted metabolizer phenotypes. No global comparisons showed significant differences. Significant post-hoc differences when compared to European reference population are indicated as following: * (p < 0.05).

**Figure 5.**
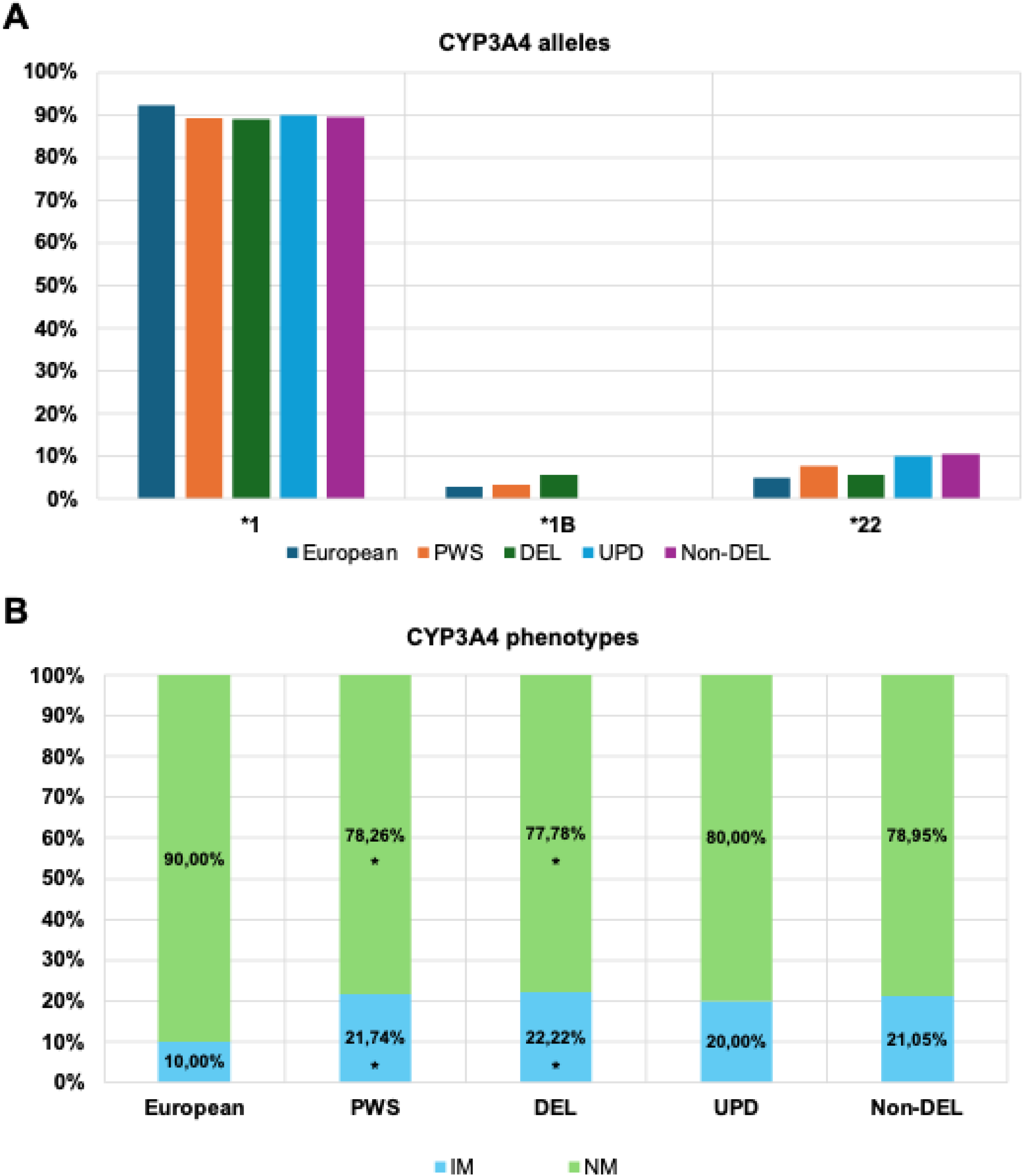
CYP3A4 allelic and predicted metabolizer phenotype distributions across study groups. (A) Allele frequency (%) in the European reference population and in the PWS cohort, including genetic subtypes. No global comparisons, nor post-hoc comparisons showed significant differences. (B) Distribution of predicted metabolizer phenotypes. Global distribution comparisons showed significant differences in European vs. PWS (p < 0.05) and European vs. DEL (p < 0.05). Significant post-hoc differences when compared to European reference population are indicated as following: * (p < 0.05).

Post-hoc analyses identified consistent enrichment of alleles CYP2B6*19 (p < 0.001; q < 0.01) and CYP2D6*10 (p < 0.001; q < 0.01) in the PWS cohort, both alleles related to a lower functionality. No other allele reached statistical significance (Figures 1A-5A; Supplementary Table 2).

#### 3.1.2. Predicted metabolizer phenotypes

Predicted metabolizer phenotype distributions revealed significant differences only for gene CYP3A4 (p = 0,02). After FDR correction, no significant differences were reported (Figures 1B-5B; Supplementary Table 1).

Post-hoc analyses showed significant enrichment of CYP3A4 IM (p = 0.02, q = 0.02) in PWS, together with a significant depletion of CYP3A4 NM (p = 0.02; q = 0.02). No other significant differences were reported (Figures 1B-5B; Supplementary Table 3).

### 3.2. European reference population vs PWS genetic subtypes

#### 3.2.1. European reference population vs DEL genetic subtype

In comparisons between European controls and DEL patients, no global allelic distribution nor post-hoc allelic analyses showed statistically significant differences (Figures 1A-5A; Supplementary Tables 1&4).

Phenotypic distribution analyses reported only a significant difference in CYP3A4 (p = 0.04) but did not remain significant after FDR correction (Figures 1B-5B; Supplementary Table 1). Post-hoc analyses showed significant enrichment of CYP3A4 IM (p = 0.04; q = 0.04) together with a significant depletion of CYP3A4 NM (p = 0.04; q = 0.04) in PWS, with no other significant differences reported (Figures 1B-5B; Supplementary Table 5).

#### 3.2.2. European reference population vs UPD genetic subtype

When comparing European controls with UPD patients, significant differences were observed in CYP2B6 (p < 0.001) and CYP2C9 (p = 0.02) global distributions, with CYP2C19 showing a borderline significant value (p = 0.06). After FDR correction, CYP2B6 (q = 0.01) was the only one that remained significant (Figures 1A-5A; Supplementary Table 1).

Post-hoc alllelic analyses showed significant enrichment of alleles CYP2B6*19 (p < 0.001), CYP2C19*3 (p = 0.04), CYP2C9*11 (p = 0.04) and CYP2D6*10 (p = 0.01) in UPD cohort. After FDR correction, only CYP2B6*19 (q = 0.001) remained significant (Figures 1A-5A; Supplementary Table 6).

These allelic differences, however, did not translate into phenotypic differences, with no significant differences in global distributions nor in post-hoc analyses reported (Figures 1B-5B; Supplementary Tables 1&7).

#### 3.2.3. European reference population vs Non-DEL genetic subtypes

Comparisons between European controls and Non-DEL patients revealed significant different allelic distributions in CYP2B6 (p = 0.001) and CYP2C19 (p = 0.04), with only CYP2B6 (q = 0.01) remaining significant after FDR correction (Figures 1A-5A; Supplementary Table 1).

Post-hoc allelic comparisons showed significant increase in CYP2B6*19 (p < 0.001; q = 0.001) and CYP2D6*10 (p < 0.001; q < 0.01) alleles in Non-DEL cohort. Alleles CYP2C19*3 (p = 0.05) and CYP2C9*11 (p = 0.05) did not reach statistical significance but remained near the threshold. No other significant differences were reported (Figures 1A-5A; Supplementary Table 8).

Global comparison of predicted metabolizer phenotypes distributions did not show any significant differences. However, post-hoc analyses showed a significant enrichment of CYP2D6 IM (p = 0.03) in the Non-DEL cohort, despite not remaining significant after FDR correction. No other significant differences were observed (Figures 1B-5B; Supplementary Tables 1&9).

### 3.3. Within-PWS comparisons by genetic subtype

Comparisons between DEL and UPD subgroups did not show any statistically significant difference, nor in global comparisons, nor in post-hoc comparisons. The same happened for comparisons between DEL and Non-Del subgroups (Figures 1-5; Supplementary Tables 1&10-13).

## 4. Discussion

PWS is characterized by hyperphagia which leads to obesity in many cases, hypotonia, respiratory difficulties, growth hormone deficiency, hypogonadotropic hypogonadism, mild cognitive impairment, along with a severe, disruptive and chronic behavioral profile that typically requires pharmacological management. Nevertheless, this approach has shown significant limitations, with common atypical responses and risk of potential severe side effects to standard dosages of psychiatric medications when used in these patients (Soni et al., 2007; Bonnot et al., 2016; Forster et al., 2021).

All these adverse effects may be explained, at least partially, by the differences observed in the pharmacogenetic profile of people with PWS. Our analysis showed differences pointing towards a potential lower metabolic capacity of psychiatric drugs in this population for all five genes studied. Main allelic differences were the increased prevalence of alleles CYP2B6*19 and CYP2D6*10, which are both associated with a decreased metabolic activity. When analyzing predicted phenotypes, the main difference arose in CYP3A4, with a significant bias towards IM, and the consequent decrease in NM. These lower metabolic capacities may induce higher drug concentrations in blood while administering standard doses of psychiatric drugs, and the subsequent adverse events.

Among all genes included, CYP2D6 emerges as the main protagonist, with high-grade evidence associations between many psychiatric medications and this enzyme. In ClinPGx, level 1A (the highest) evidence associations are described for imipramine, nortriptyline, atomoxetine, amitriptyline, desipramine, clomipramine, vortioxetine, paroxetine, fluvoxamine, haloperidol, trimipramine, venlafaxine, aripiprazole and risperidone. Our cohort displayed a lower metabolic profile for CYP2D6, meaning that a worse metabolism of all these drugs, and the consequent higher risk of adverse events at standard dosages, might be expected. This is highly relevant, since aripiprazole and risperidone are quite commonly prescribed in PWS. For aripiprazole, a report of 10 adults with PWS receiving this medication for temper-outburst management showed a 60% prevalence of adverse events, specifically, excessive daytime sleepiness, leading to medication discontinuation in 4 of the 6 patients affected (Deest et al., 2022). For fluoxetine, a common psychotropic medication in PWS, there is no high-grade evidence association with any gene, but still is mainly metabolized by CYP2D6, CYP2C9 and CYP3A4; and most importantly, is a strong inhibitor of CYP2D6, meaning that it could decrease even more the metabolic activity of this enzyme. Also, there have been cases of fluoxetine-induced psychosis (Carpenter et al., 1998), even a case of a pediatric female with PWS (Hergüner and Motavalli Mukaddes, 2007). With psychosis being prevalent in PWS, administration of fluoxetine, which is frequently prescribed, may need to be more carefully regulated, even more in a population that has an increased baseline risk of developing psychosis (Soni et al., 2007).

For CYP2B6, whose activity seemed also to be reduced in our PWS cohort, a high-grade evidence association in ClinPGx has been described with sertraline, suggesting that lower metabolic rates for CYP2B6 are related to a lower sertraline metabolism. Forster showed that people with PWS receiving sertraline may be at risk of suffering from mood and behavioral activation (MBA) induced by this psychotropic drug (Forster, 2025). Other ClinPGx high-grade evidence associations include CYP3A4 with quetiapine; and CYP2C19 with citalopram, escitalopram and sertraline (Milosavljević et al., 2021).

Despite not founding any significant difference between PWS subgroups, CYP2C19 emerged as an interesting gene. CYP2C19 phenotypic landscape appears to tend to a lower metabolic activity in PWS. Nevertheless, when stratifying for PWS subgroup, there were opposite situations. DEL subtype showed an increased prevalence of faster metabolizing phenotypes, while UPD and Non-DEL groups showed clear prevalence of lower metabolizing phenotypes, despite not reaching significant differences in global distributions. Taking together that PWS subtypes differ in their psychiatric comorbidities, potential differences in metabolic capabilities should be analyzed when psychotropic drugs should be used.

Our results are coherent with Forster et al. work in a cohort of north American patients with PWS (n=34), the only pharmacogenetic characterization of patients with this syndrome published to date (Forster et al., 2021). They reported a similar shift towards CYP3A4 IM frequency compared to our cohort, together with tendencies towards lower metabolic capacity. Moreover, they also reported tendencies towards a slower metabolic capacibility for all genes included in our study. Despite geographical and selection differences (our cohort was randomly selected and that of Forster et al was of individuals with previous psychotropic drug failure), both cohorts displayed similar pharmacogenetic characteristics. These results reinforce our hypothesis that individuals with PWS show impaired metabolic capacity and, thus, may need different guidelines for psychiatric drug prescription.

On the other hand, there are other factors that modulate drug metabolism. Drugs may act as potential substrates, inducers and/or inhibitors of enzymes of the CYP450 family (Deodhar et al., 2020). Thus, concurrent administration of different medications that happen to interact with the same CYP450 enzyme can lead to atypical responses. Also, growth hormone (GH) replacement therapy, which is a standard of care in PWS (Casamitjana et al., 2022), may alter CYP3A4 activity. CYP3A4 activity differs in males and females, due to its modulation by GH secretion. GH secretion follows a pulsatile pattern in male infants, and a more continuous pattern in female infants. GH secretion patterns regulate CYP3A4 expression in the liver through transcription factor STAT5b, which inhibits liver CYP3A4 expression. In males, pulsatile secretion, in bursts, sensitizes STAT5b, so that its activation is faster in response to GH signaling. In the case of GH deficient infants, such as in PWS, patients exhibit increased levels of CYP3A4 expression, since there is no inhibition. However, when administering GH replacement therapy, since the dosage mimics a continuous pattern, it has a stronger effect in male patients, who are more sensitized to GH. Thus, male infants with GH replacement therapy achieve normal CYP3A4 hepatic expression levels, whilst female infants may still exhibit an increased expression (Sinués et al., 2004). And finally, body weight also plays an important role in drug metabolism, distribution and clinical responses. Obesity, typically present in PWS, can affect volume of distribution, clearance rates and drug sensitivity, potentially leading to atypical responses to drugs at standard doses (Hanley et al., 2010). Thus, all these potential interfering factors should be considered alongside pharmacogenetics when selecting and dosing medications.

### Limitations and Strengths

The main limitation of this study is the small sample size of the PWS cohort, which limits the statistical power for the comparisons. More limitations include the lack of reference data for genes CYP1A2 and CYP3A4, which are not available in ClinPGx. Another limitation is the role of allele CYP3A4*1B, whose phenotypic activity remains unconclusive. In this article, we considered it had a decreased activity, mainly because this work followed the line of Forster et al. who considered this assignation to this allele (Forster et al., 2021). Finally, phenotypic activity degrees assigned to individuals in this study are predicted and, since there are many other factors present in PWS that may alter metabolic activity of CYP450 genes, the real metabolic capacity of people living with PWS should also be addressed through metabolite quantification methods to confirm if predictions are accurate or not.

## 5. Conclusions

In conclusion, our PWS cohort exhibited tendencies towards a lower metabolic capacity of genes CYP2B6, CYP2C9, CYP2C19, CYP2D6 and CYP3A4, which may constitute a potential explanation for the atypical responses towards standard doses of psychiatric medications observed in individuals with PWS.

Future studies shall increase sample size and potentially include a reference sample of individuals without PWS from the same geographical locations to confirm these findings and generate better and more adapted prescription guidelines of psychiatric medications for people with PWS.

## Data Availability

All data produced in the present work are contained in the manuscript.

