## Supplementary Tables for "Pharmacogenetic Characterization of Cytochrome P450 Genes involved in Psychotropic Medication Metabolism in a Cohort of Patients with Prader-Willi Syndrome"

| **Populations** | **Comparison** | **Gene** | **p-value** | **q-value (FDR-corrected)** |
| --- | --- | --- | --- | --- |
| European vs PWS | Allele | CYP2B6 | **0,006571993** | 0,065719934 |
|  |  | CYP2C19 | 0,066955933 | 0,251084749 |
|  |  | CYP2C9 | 0,196180804 | 0,452724932 |
|  |  | CYP2D6 | 0,052088948 | 0,251084749 |
|  |  | CYP3A4 | 0,384614615 | 0,588749411 |
|  | Phenotype | CYP2B6 | 0,836564163 | 1 |
|  |  | CYP2C19 | 0,414034586 | 0,797314497 |
|  |  | CYP2C9 | 0,443257557 | 0,797314497 |
|  |  | CYP2D6 | 0,237328763 | 0,797314497 |
|  |  | CYP3A4 | **0,021882925** | 0,65648776 |
| European vs DEL | Allele | CYP2B6 | 0,906308094 | 0,971044386 |
|  |  | CYP2C19 | 0,137634862 | 0,375367806 |
|  |  | CYP2C9 | 0,859016141 | 0,954462379 |
|  |  | CYP2D6 | 0,457927542 | 0,598564619 |
|  |  | CYP3A4 | 0,315850684 | 0,588749411 |
|  | Phenotype | CYP2B6 | 0,725199275 | 1 |
|  |  | CYP2C19 | 0,225036775 | 0,797314497 |
|  |  | CYP2C9 | 0,782184218 | 1 |
|  |  | CYP2D6 | 0,385601614 | 0,797314497 |
|  |  | CYP3A4 | **0,047122386** | 0,706835784 |
| European vs UPD | Allele | CYP2B6 | **0,000798999** | **0,017039983** |
|  |  | CYP2C19 | 0,061464939 | 0,251084749 |
|  |  | CYP2C9 | **0,027478973** | 0,206092294 |
|  |  | CYP2D6 | 0,384196616 | 0,588749411 |
|  |  | CYP3A4 | 0,353887646 | 0,588749411 |
|  | Phenotype | CYP2B6 | 0,588798411 | 0,981330685 |
|  |  | CYP2C19 | 0,426333574 | 0,797314497 |
|  |  | CYP2C9 | 1 | 1 |
|  |  | CYP2D6 | 0,861246139 | 1 |
|  |  | CYP3A4 | 0,184145997 | 0,797314497 |
| European vs Non-DEL | Allele | CYP2B6 | **0,001135999** | **0,017039983** |
|  |  | CYP2C19 | **0,048356952** | 0,251084749 |
|  |  | CYP2C9 | 0,104381896 | 0,330878669 |
|  |  | CYP2D6 | 0,11029289 | 0,330878669 |
|  |  | CYP3A4 | 0,167955832 | 0,41988958 |
|  | Phenotype | CYP2B6 | 0,751387249 | 1 |
|  |  | CYP2C19 | 0,315834684 | 0,797314497 |
|  |  | CYP2C9 | 0,667382333 | 1 |
|  |  | CYP2D6 | 0,321633678 | 0,797314497 |
|  |  | CYP3A4 | 0,115080294 | 0,797314497 |
| DEL vs UPD | Allele | CYP2B6 | 0,50953949 | 0,61361339 |
|  |  | CYP2C19 | 0,42379458 | 0,57790169 |
|  |  | CYP2C9 | 0,39348761 | 0,56212515 |
|  |  | CYP2D6 | 0,996981 | 0,996981 |
|  |  | CYP3A4 | 0,45936654 | 0,59917375 |
|  | Phenotype | CYP2B6 | 0,35676364 | 0,79669509 |
|  |  | CYP2C19 | 0,42453558 | 0,79669509 |
|  |  | CYP2C9 | 0,74597525 | 1 |
|  |  | CYP2D6 | 0,92057408 | 1 |
|  |  | CYP3A4 | 1 | 1 |
| DEL vs Non-DEL | Allele | CYP2B6 | 0,51134449 | 0,61361339 |
|  |  | CYP2C19 | 0,22141678 | 0,47446453 |
|  |  | CYP2C9 | 0,82833417 | 0,95397905 |
|  |  | CYP2D6 | 0,96177704 | 0,99494176 |
|  |  | CYP3A4 | 0,28053872 | 0,56107744 |
|  | Phenotype | CYP2B6 | 0,3977596 | 0,79669509 |
|  |  | CYP2C19 | 0,21708778 | 0,79669509 |
|  |  | CYP2C9 | 1 | 1 |
|  |  | CYP2D6 | 0,45146055 | 0,79669509 |
|  |  | CYP3A4 | 1 | 1 |

Supplementary Table 1. Global analysis comparisons of frequency distributions between populations. All comparisons were performed using Fisher exact test with Monte Carlo simulations (10⁶ iterations), with significance shown as p-values. False Discovery Rate Benjamini-Hochberg corrections were applied (q-values). Significant p- and q-values are shown in bold.

| **Gene** | **Allele** | **European Frequency** | **PWS Frequency** | **Delta Frequency** | **p-value** | **q-value (FDR-corrected)** |
| --- | --- | --- | --- | --- | --- | --- |
| CYP2B6 | *1 | 0,52019352 | 0,53333333 | -0,01313982 | 0,83340069 | 1 |
|  | *19 | 0 | 0,01111111 | -0,01111111 | **0,00068522** | **0,004796528** |
|  | *2 | 0,05215801 | 0,05555556 | -0,00339754 | 0,81115907 | 1 |
|  | *22 | 0,01484134 | 0,01111111 | 0,00373023 | 1 | 1 |
|  | *4 | 0,04335835 | 0,03333333 | 0,01002501 | 1 | 1 |
|  | *5 | 0,12244105 | 0,08888889 | 0,03355216 | 0,42080379 | 1 |
|  | *6 | 0,24700773 | 0,26666667 | -0,01965893 | 0,62719022 | 1 |
| CYP2C19 | *1 | 0,63055835 | 0,58888889 | 0,04166946 | 0,44477958 | 0,555974479 |
|  | *17 | 0,21730333 | 0,2 | 0,01730333 | 0,79834687 | 0,798346873 |
|  | *2 | 0,14812655 | 0,18888889 | -0,04076234 | 0,29699171 | 0,49498619 |
|  | *3 | 0,00163 | 0,01111111 | -0,00948111 | 0,13705662 | 0,484009607 |
|  | *4 | 0,00238177 | 0,01111111 | -0,00872934 | 0,19360384 | 0,484009607 |
| CYP2C9 | *1 | 0,79494377 | 0,78888889 | 0,00605488 | 0,89604441 | 0,896044411 |
|  | *11 | 0,00165326 | 0,01111111 | -0,00945785 | 0,13959442 | 0,558377698 |
|  | *2 | 0,12764565 | 0,14444444 | -0,01679879 | 0,63436981 | 0,896044411 |
|  | *3 | 0,07575731 | 0,05555556 | 0,02020176 | 0,6876708 | 0,896044411 |
| CYP2D6 | *1 | 0,3257619 | 0,23404255 | 0,09171935 | 0,06129001 | 0,367740046 |
|  | *10 | 0,01796423 | 0,08510638 | -0,06714215 | **0,00031** | **0,003719953** |
|  | *2 | 0,21186552 | 0,27659574 | -0,06473022 | 0,12992041 | 0,519681642 |
|  | *3 | 0,01820129 | 0,0212766 | -0,0030753 | 0,69002495 | 1 |
|  | *34 | 0,01228346 | 0,0106383 | 0,00164516 | 1 | 1 |
|  | *4 | 0,21127725 | 0,19148936 | 0,01978789 | 0,70583648 | 1 |
|  | *41 | 0,10558155 | 0,09574468 | 0,00983687 | 0,8676544 | 1 |
|  | *5 | 0,0336983 | 0,03191489 | 0,0017834 | 1 | 1 |
|  | *6 | 0,01280149 | 0 | 0,01280149 | 0,63673558 | 1 |
|  | *9 | 0,03147691 | 0,03191489 | -0,00043798 | 0,77157505 | 1 |
|  | Dup1 | 0,00936844 | 0,0106383 | -0,00126986 | 0,5873877 | 1 |
|  | Dup2 | 0,00971965 | 0,0106383 | -0,00091865 | 0,60090855 | 1 |
| CYP3A4 | *1 | 0,92250373 | 0,89130435 | 0,03119938 | 0,24224597 | 0,363368958 |
|  | *1B | 0,02779758 | 0,0326087 | -0,00481112 | 0,74415546 | 0,744155458 |
|  | *22 | 0,0496987 | 0,07608696 | -0,02638826 | 0,2268863 | 0,363368958 |

Supplementary Table 2. Post-hoc allelic frequency comparisons between European and PWS populations. All comparisons were performed using Fisher exact test, with significance shown as p-values. False Discovery Rate Benjamini-Hochberg corrections were applied (q-values). Significant p- and q-values are shown in bold.

| **Gene** | **Phenotype** | **European Frequency** | **PWS Frequency** | **Delta Frequency** | **p-value** | **q-value (FDR-corrected)** |
| --- | --- | --- | --- | --- | --- | --- |
| CYP2B6 | UM | 0,00169608 | 0 | 0,00169608 | 1 | 1 |
|  | RM | 0,05359915 | 0,02222222 | 0,03137693 | 0,51760396 | 0,97921483 |
|  | NM | 0,4294975 | 0,46666667 | -0,03716917 | 0,65280989 | 0,97921483 |
|  | IM | 0,37950612 | 0,42222222 | -0,0427161 | 0,5433442 | 0,97921483 |
|  | PM | 0,07429713 | 0,06666667 | 0,00763046 | 1 | 1 |
|  | Other | 0,06140401 | 0,02222222 | 0,03918179 | 0,52568991 | 0,97921483 |
| CYP2C19 | UM | 0,04641832 | 0,02222222 | 0,0241961 | 0,72359434 | 1 |
|  | RM | 0,27118219 | 0,26666667 | 0,00451552 | 1 | 1 |
|  | NM | 0,39611602 | 0,35555556 | 0,04056046 | 0,64894529 | 1 |
|  | IM | 0,26108217 | 0,28888889 | -0,02780672 | 0,73411192 | 1 |
|  | PM | 0,02387785 | 0,06666667 | -0,04278881 | 0,09245201 | 0,55471206 |
|  | Other | 0,00132345 | 0 | 0,00132345 | 1 | 1 |
| CYP2C9 | NM | 0,62846053 | 0,57777778 | 0,05068276 | 0,53743537 | 0,8424525 |
|  | IM | 0,34526345 | 0,42222222 | -0,07695878 | 0,27625855 | 0,8424525 |
|  | PM | 0,02569211 | 0 | 0,02569211 | 0,63183938 | 0,8424525 |
|  | Other | 0,00058391 | 0 | 0,00058391 | 1 | 1 |
| CYP2D6 | UM | 0,02330619 | 0,04255319 | -0,019247 | 0,2999335 | 0,52819571 |
|  | NM | 0,49190352 | 0,44680851 | 0,04509501 | 0,56236967 | 0,56236967 |
|  | IM | 0,38253188 | 0,4893617 | -0,10682982 | 0,1358364 | 0,52819571 |
|  | PM | 0,06501767 | 0,0212766 | 0,04374107 | 0,36954813 | 0,52819571 |
|  | Other | 0,03724074 | 0 | 0,03724074 | 0,42255657 | 0,52819571 |
| CYP3A4 | NM | 0,89999352 | 0,7826087 | 0,11738482 | **0,02188293** | **0,02188293** |
|  | IM | 0,10000648 | 0,2173913 | -0,11738482 | **0,02188293** | **0,02188293** |

Supplementary Table 3. Post-hoc phenotypic frequency comparisons between European and PWS populations. All comparisons were performed using Fisher exact test, with significance shown as p-values. False Discovery Rate Benjamini-Hochberg corrections were applied (q-values). Significant p- and q-values are shown in bold.

| **Gene** | **Allele** | **European Frequency** | **PWS Frequency** | **Delta Frequency** | **p-value** | **q-value (FDR-corrected)** |
| --- | --- | --- | --- | --- | --- | --- |
| CYP2B6 | *1 | 0,52019352 | 0,55555556 | -0,035362 | 0,68331368 | 1 |
|  | *19 | 0 | 0 | 0 | 1 | 1 |
|  | *2 | 0,05215801 | 0,03703704 | 0,01512098 | 1 | 1 |
|  | *22 | 0,01484134 | 0 | 0,01484134 | 1 | 1 |
|  | *4 | 0,04335835 | 0,01851852 | 0,02483983 | 0,73155414 | 1 |
|  | *5 | 0,12244105 | 0,09259259 | 0,02984846 | 0,67750393 | 1 |
|  | *6 | 0,24700773 | 0,2962963 | -0,0492886 | 0,42974567 | 1 |
| CYP2C19 | *1 | 0,63055835 | 0,53703704 | 0,09352131 | 0,16041092 | 0,4010273 |
|  | *17 | 0,21730333 | 0,25925926 | -0,0419559 | 0,50830125 | 0,63537656 |
|  | *2 | 0,14812655 | 0,18518519 | -0,0370586 | 0,44224132 | 0,63537656 |
|  | *3 | 0,00163 | 0 | 0,00163 | 1 | 1 |
|  | *4 | 0,00238177 | 0,01851852 | -0,0161367 | 0,1211354 | 0,4010273 |
| CYP2C9 | *1 | 0,79494377 | 0,7962963 | -0,0013525 | 1 | 1 |
|  | *11 | 0,00165326 | 0 | 0,00165326 | 1 | 1 |
|  | *2 | 0,12764565 | 0,14814815 | -0,0205025 | 0,68137374 | 1 |
|  | *3 | 0,07575731 | 0,05555556 | 0,02020176 | 0,79712004 | 1 |
| CYP2D6 | *1 | 0,3257619 | 0,25 | 0,0757619 | 0,25576025 | 0,84300577 |
|  | *10 | 0,01796423 | 0,05357143 | -0,0356072 | 0,07986214 | 0,84300577 |
|  | *2 | 0,21186552 | 0,28571429 | -0,0738488 | 0,189954 | 0,84300577 |
|  | *3 | 0,01820129 | 0,01785714 | 0,00034415 | 1 | 1 |
|  | *34 | 0,01228346 | 0,01785714 | -0,0055737 | 0,4996547 | 0,85655092 |
|  | *4 | 0,21127725 | 0,16071429 | 0,05056297 | 0,41566119 | 0,84300577 |
|  | *41 | 0,10558155 | 0,10714286 | -0,0015613 | 1 | 1 |
|  | *5 | 0,0336983 | 0,03571429 | -0,002016 | 0,71358387 | 0,95144516 |
|  | *6 | 0,01280149 | 0 | 0,01280149 | 1 | 1 |
|  | *9 | 0,03147691 | 0,03571429 | -0,0042374 | 0,69644879 | 0,95144516 |
|  | Dup1 | 0,00936844 | 0,01785714 | -0,0084887 | 0,40990366 | 0,84300577 |
|  | Dup2 | 0,00971965 | 0,01785714 | -0,0081375 | 0,42150289 | 0,84300577 |
| CYP3A4 | *1 | 0,92250373 | 0,88888889 | 0,03361484 | 0,30986656 | 0,46479984 |
|  | *1B | 0,02779758 | 0,05555556 | -0,027758 | 0,18973086 | 0,46479984 |
|  | *22 | 0,0496987 | 0,05555556 | -0,0058569 | 0,75045474 | 0,75045474 |

Supplementary Table 4. Post-hoc allelic frequency comparisons between European and DEL populations. All comparisons were performed using Fisher exact test, with significance shown as p-values. False Discovery Rate Benjamini-Hochberg corrections were applied (q-values). Significant p- and q-values are shown in bold.

| **Gene** | **Phenotype** | **European Frequency** | **PWS Frequency** | **Delta Frequency** | **p-value** | **q-value (FDR-corrected)** |
| --- | --- | --- | --- | --- | --- | --- |
| CYP2B6 | UM | 0,00169608 | 0 | 0,00169608 | 1 | 1 |
|  | RM | 0,05359915 | 0,03703704 | 0,01656211 | 1 | 1 |
|  | NM | 0,4294975 | 0,48148148 | -0,051984 | 0,69812691 | 1 |
|  | IM | 0,37950612 | 0,37037037 | 0,00913575 | 1 | 1 |
|  | PM | 0,07429713 | 0,11111111 | -0,036814 | 0,44926157 | 1 |
|  | Other | 0,06140401 | 0 | 0,06140401 | 0,40927611 | 1 |
| CYP2C19 | UM | 0,04641832 | 0,03703704 | 0,00938129 | 1 | 1 |
|  | RM | 0,27118219 | 0,37037037 | -0,0991882 | 0,27840067 | 0,55680134 |
|  | NM | 0,39611602 | 0,25925926 | 0,13685676 | 0,1708898 | 0,51266939 |
|  | IM | 0,26108217 | 0,25925926 | 0,00182291 | 1 | 1 |
|  | PM | 0,02387785 | 0,07407407 | -0,0501962 | 0,135491 | 0,51266939 |
|  | Other | 0,00132345 | 0 | 0,00132345 | 1 | 1 |
| CYP2C9 | NM | 0,62846053 | 0,59259259 | 0,03586794 | 0,69463757 | 1 |
|  | IM | 0,34526345 | 0,40740741 | -0,062144 | 0,54497965 | 1 |
|  | PM | 0,02569211 | 0 | 0,02569211 | 1 | 1 |
|  | Other | 0,00058391 | 0 | 0,00058391 | 1 | 1 |
| CYP2D6 | UM | 0,02330619 | 0,07142857 | -0,0481224 | 0,13824424 | 0,69122119 |
|  | NM | 0,49190352 | 0,46428571 | 0,0276178 | 0,8510715 | 1 |
|  | IM | 0,38253188 | 0,42857143 | -0,0460395 | 0,69810048 | 1 |
|  | PM | 0,06501767 | 0,03571429 | 0,02930338 | 1 | 1 |
|  | Other | 0,03724074 | 0 | 0,03724074 | 0,62567212 | 1 |
| CYP3A4 | NM | 0,89999352 | 0,77777778 | 0,12221574 | **0,04712239** | **0,04712239** |
|  | IM | 0,10000648 | 0,22222222 | -0,1222157 | **0,04712239** | **0,04712239** |

Supplementary Table 5. Post-hoc phenotypic frequency comparisons between European and DEL populations. All comparisons were performed using Fisher exact test, with significance shown as p-values. False Discovery Rate Benjamini-Hochberg corrections were applied (q-values). Significant p- and q-values are shown in bold.

| **Gene** | **Allele** | **European Frequency** | **PWS Frequency** | **Delta Frequency** | **p-value** | **q-value (FDR-corrected)** |
| --- | --- | --- | --- | --- | --- | --- |
| CYP2B6 | *1 | 0,52019352 | 0,53333333 | -0,0131398 | 1 | 1 |
|  | *19 | 0 | 0,03333333 | -0,0333333 | **0,00022851** | **0,00159957** |
|  | *2 | 0,05215801 | 0,06666667 | -0,0145087 | 0,6690288 | 1 |
|  | *22 | 0,01484134 | 0 | 0,01484134 | 1 | 1 |
|  | *4 | 0,04335835 | 0,06666667 | -0,0233083 | 0,37585076 | 0,87698511 |
|  | *5 | 0,12244105 | 0,03333333 | 0,08910772 | 0,16927908 | 0,59247676 |
|  | *6 | 0,24700773 | 0,26666667 | -0,0196589 | 0,83253162 | 1 |
| CYP2C19 | *1 | 0,63055835 | 0,63333333 | -0,002775 | 1 | 1 |
|  | *17 | 0,21730333 | 0,13333333 | 0,08396999 | 0,37523678 | 0,72848296 |
|  | *2 | 0,14812655 | 0,2 | -0,0518734 | 0,43708978 | 0,72848296 |
|  | *3 | 0,00163 | 0,03333333 | -0,0317033 | **0,04795765** | 0,23978826 |
|  | *4 | 0,00238177 | 0 | 0,00238177 | 1 | 1 |
| CYP2C9 | *1 | 0,79494377 | 0,83333333 | -0,0383896 | 0,82091833 | 0,82091833 |
|  | *11 | 0,00165326 | 0,03333333 | -0,0316801 | **0,04890662** | 0,19562649 |
|  | *2 | 0,12764565 | 0,13333333 | -0,0056877 | 0,78849919 | 0,82091833 |
|  | *3 | 0,07575731 | 0 | 0,07575731 | 0,16669424 | 0,33338848 |
| CYP2D6 | *1 | 0,3257619 | 0,23333333 | 0,09242857 | 0,33378408 | 1 |
|  | *10 | 0,01796423 | 0,1 | -0,0820358 | **0,01642855** | 0,19714257 |
|  | *2 | 0,21186552 | 0,3 | -0,0881345 | 0,26197743 | 1 |
|  | *3 | 0,01820129 | 0,03333333 | -0,015132 | 0,42377775 | 1 |
|  | *34 | 0,01228346 | 0 | 0,01228346 | 1 | 1 |
|  | *4 | 0,21127725 | 0,2 | 0,01127725 | 1 | 1 |
|  | *41 | 0,10558155 | 0,06666667 | 0,03891489 | 0,76506257 | 1 |
|  | *5 | 0,0336983 | 0,03333333 | 0,00036496 | 1 | 1 |
|  | *6 | 0,01280149 | 0 | 0,01280149 | 1 | 1 |
|  | *9 | 0,03147691 | 0,03333333 | -0,0018564 | 0,61696465 | 1 |
|  | Dup1 | 0,00936844 | 0 | 0,00936844 | 1 | 1 |
|  | Dup2 | 0,00971965 | 0 | 0,00971965 | 1 | 1 |
| CYP3A4 | *1 | 0,92250373 | 0,9 | 0,02250373 | 0,50289323 | 0,75433984 |
|  | *1B | 0,02779758 | 0 | 0,02779758 | 1 | 1 |
|  | *22 | 0,0496987 | 0,1 | -0,0503013 | 0,18558892 | 0,55676675 |

Supplementary Table 6. Post-hoc allelic frequency comparisons between European and UPD populations. All comparisons were performed using Fisher exact test, with significance shown as p-values. False Discovery Rate Benjamini-Hochberg corrections were applied (q-values). Significant p- and q-values are shown in bold.

| **Gene** | **Phenotype** | **European Frequency** | **PWS Frequency** | **Delta Frequency** | **p-value** | **q-value (FDR-corrected)** |
| --- | --- | --- | --- | --- | --- | --- |
| CYP2B6 | UM | 0,00169608 | 0 | 0,00169608 | 1 | 1 |
|  | RM | 0,05359915 | 0 | 0,05359915 | 1 | 1 |
|  | NM | 0,4294975 | 0,4 | 0,0294975 | 1 | 1 |
|  | IM | 0,37950612 | 0,6 | -0,2204939 | 0,10810662 | 0,64863969 |
|  | PM | 0,07429713 | 0 | 0,07429713 | 0,62180202 | 1 |
|  | Other | 0,06140401 | 0 | 0,06140401 | 1 | 1 |
| CYP2C19 | UM | 0,04641832 | 0 | 0,04641832 | 1 | 1 |
|  | RM | 0,27118219 | 0,13333333 | 0,13784886 | 0,38240009 | 0,90688144 |
|  | NM | 0,39611602 | 0,46666667 | -0,0705506 | 0,60458763 | 0,90688144 |
|  | IM | 0,26108217 | 0,33333333 | -0,0722512 | 0,55803849 | 0,90688144 |
|  | PM | 0,02387785 | 0,06666667 | -0,0427888 | 0,3041954 | 0,90688144 |
|  | Other | 0,00132345 | 0 | 0,00132345 | 1 | 1 |
| CYP2C9 | NM | 0,62846053 | 0,66666667 | -0,0382061 | 1 | 1 |
|  | IM | 0,34526345 | 0,33333333 | 0,01193011 | 1 | 1 |
|  | PM | 0,02569211 | 0 | 0,02569211 | 1 | 1 |
|  | Other | 0,00058391 | 0 | 0,00058391 | 1 | 1 |
| CYP2D6 | UM | 0,02330619 | 0 | 0,02330619 | 1 | 1 |
|  | NM | 0,49190352 | 0,46666667 | 0,02523685 | 1 | 1 |
|  | IM | 0,38253188 | 0,53333333 | -0,1508015 | 0,28879831 | 1 |
|  | PM | 0,06501767 | 0 | 0,06501767 | 0,61944377 | 1 |
|  | Other | 0,03724074 | 0 | 0,03724074 | 1 | 1 |
| CYP3A4 | NM | 0,89999352 | 0,8 | 0,09999352 | 0,184146 | 0,184146 |
|  | IM | 0,10000648 | 0,2 | -0,0999935 | 0,184146 | 0,184146 |

Supplementary Table 7. Post-hoc phenotypic frequency comparisons between European and UPD populations. All comparisons were performed using Fisher exact test, with significance shown as p-values. False Discovery Rate Benjamini-Hochberg corrections were applied (q-values). Significant p- and q-values are shown in bold.

| **Gene** | **Allele** | **European Frequency** | **PWS Frequency** | **Delta Frequency** | **p-value** | **q-value (FDR-corrected)** |
| --- | --- | --- | --- | --- | --- | --- |
| CYP2B6 | *1 | 0,52019352 | 0,5 | 0,02019352 | 0,86826959 | 0,86826959 |
|  | *19 | 0 | 0,02777778 | -0,0277778 | **0,0002742** | **0,0019194** |
|  | *2 | 0,05215801 | 0,08333333 | -0,0311753 | 0,43466512 | 0,86826959 |
|  | *22 | 0,01484134 | 0,02777778 | -0,0129364 | 0,41637382 | 0,86826959 |
|  | *4 | 0,04335835 | 0,05555556 | -0,0121972 | 0,66916827 | 0,86826959 |
|  | *5 | 0,12244105 | 0,08333333 | 0,03910772 | 0,61638429 | 0,86826959 |
|  | *6 | 0,24700773 | 0,22222222 | 0,02478551 | 0,84808998 | 0,86826959 |
| CYP2C19 | *1 | 0,63055835 | 0,66666667 | -0,0361083 | 0,73183172 | 0,91478965 |
|  | *17 | 0,21730333 | 0,11111111 | 0,10619222 | 0,15636019 | 0,39090047 |
|  | *2 | 0,14812655 | 0,19444444 | -0,0463179 | 0,47779089 | 0,79631815 |
|  | *3 | 0,00163 | 0,02777778 | -0,0261478 | 0,05726841 | 0,28634204 |
|  | *4 | 0,00238177 | 0 | 0,00238177 | 1 | 1 |
| CYP2C9 | *1 | 0,79494377 | 0,77777778 | 0,01716599 | 0,8361118 | 1 |
|  | *11 | 0,00165326 | 0,02777778 | -0,0261245 | 0,05839417 | 0,23357667 |
|  | *2 | 0,12764565 | 0,13888889 | -0,0112432 | 0,8021019 | 1 |
|  | *3 | 0,07575731 | 0,05555556 | 0,02020176 | 1 | 1 |
| CYP2D6 | *1 | 0,3257619 | 0,21052632 | 0,11523558 | 0,16547655 | 0,99285929 |
|  | *10 | 0,01796423 | 0,13157895 | -0,1136147 | **0,00057626** | **0,00691512** |
|  | *2 | 0,21186552 | 0,26315789 | -0,0512924 | 0,4290008 | 1 |
|  | *3 | 0,01820129 | 0,02631579 | -0,0081145 | 0,50254006 | 1 |
|  | *34 | 0,01228346 | 0 | 0,01228346 | 1 | 1 |
|  | *4 | 0,21127725 | 0,23684211 | -0,0255649 | 0,69171684 | 1 |
|  | *41 | 0,10558155 | 0,07894737 | 0,02663419 | 0,7934834 | 1 |
|  | *5 | 0,0336983 | 0,02631579 | 0,00738251 | 1 | 1 |
|  | *6 | 0,01280149 | 0 | 0,01280149 | 1 | 1 |
|  | *9 | 0,03147691 | 0,02631579 | 0,00516112 | 1 | 1 |
|  | Dup1 | 0,00936844 | 0 | 0,00936844 | 1 | 1 |
|  | Dup2 | 0,00971965 | 0 | 0,00971965 | 1 | 1 |
| CYP3A4 | *1 | 0,92250373 | 0,89473684 | 0,02776688 | 0,53530392 | 0,62774107 |
|  | *1B | 0,02779758 | 0 | 0,02779758 | 0,62774107 | 0,62774107 |
|  | *22 | 0,0496987 | 0,10526316 | -0,0555645 | 0,11850741 | 0,35552224 |

Supplementary Table 8. Post-hoc allelic frequency comparisons between European and Non-DEL populations. All comparisons were performed using Fisher exact test, with significance shown as p-values. False Discovery Rate Benjamini-Hochberg corrections were applied (q-values). Significant p- and q-values are shown in bold.

| **Gene** | **Phenotype** | **European Frequency** | **PWS Frequency** | **Delta Frequency** | **p-value** | **q-value (FDR-corrected)** |
| --- | --- | --- | --- | --- | --- | --- |
| CYP2B6 | UM | 0,00169608 | 0 | 0,00169608 | 1 | 1 |
|  | RM | 0,05359915 | 0 | 0,05359915 | 0,62176534 | 1 |
|  | NM | 0,4294975 | 0,44444444 | -0,0149469 | 1 | 1 |
|  | IM | 0,37950612 | 0,5 | -0,1204939 | 0,33429888 | 1 |
|  | PM | 0,07429713 | 0 | 0,07429713 | 0,63996823 | 1 |
|  | Other | 0,06140401 | 0,05555556 | 0,00584846 | 1 | 1 |
| CYP2C19 | UM | 0,04641832 | 0 | 0,04641832 | 1 | 1 |
|  | RM | 0,27118219 | 0,11111111 | 0,16007108 | 0,18369748 | 0,88621891 |
|  | NM | 0,39611602 | 0,5 | -0,103884 | 0,47053756 | 0,88621891 |
|  | IM | 0,26108217 | 0,33333333 | -0,0722512 | 0,59081261 | 0,88621891 |
|  | PM | 0,02387785 | 0,05555556 | -0,0316777 | 0,35287414 | 0,88621891 |
|  | Other | 0,00132345 | 0 | 0,00132345 | 1 | 1 |
| CYP2C9 | NM | 0,62846053 | 0,55555556 | 0,07290498 | 0,62667221 | 1 |
|  | IM | 0,34526345 | 0,44444444 | -0,099181 | 0,45751924 | 1 |
|  | PM | 0,02569211 | 0 | 0,02569211 | 1 | 1 |
|  | Other | 0,00058391 | 0 | 0,00058391 | 1 | 1 |
| CYP2D6 | UM | 0,02330619 | 0 | 0,02330619 | 1 | 1 |
|  | NM | 0,49190352 | 0,36842105 | 0,12348247 | 0,36043239 | 0,90108097 |
|  | IM | 0,38253188 | 0,63157895 | -0,2490471 | **0,03268163** | 0,16340814 |
|  | PM | 0,06501767 | 0 | 0,06501767 | 0,63159351 | 1 |
|  | Other | 0,03724074 | 0 | 0,03724074 | 1 | 1 |
| CYP3A4 | NM | 0,89999352 | 0,78947368 | 0,11051984 | 0,11508029 | 0,11508029 |
|  | IM | 0,10000648 | 0,21052632 | -0,1105198 | 0,11508029 | 0,11508029 |

Supplementary Table 9. Post-hoc phenotypic frequency comparisons between European and Non-DEL populations. All comparisons were performed using Fisher exact test, with significance shown as p-values. False Discovery Rate Benjamini-Hochberg corrections were applied (q-values). Significant p- and q-values are shown in bold.

| **Gene** | **Allele** | **European Frequency** | **PWS Frequency** | **Delta Frequency** | **p-value** | **q-value (FDR-corrected)** |
| --- | --- | --- | --- | --- | --- | --- |
| CYP2B6 | *1 | 0,55555556 | 0,53333333 | 0,02222222 | 1 | 1 |
|  | *19 | 0 | 0,03333333 | -0,0333333 | 0,35714286 | 0,9655681 |
|  | *2 | 0,03703704 | 0,06666667 | -0,0296296 | 0,61434589 | 1 |
|  | *22 | 0 | 0 | 0 | 1 | 1 |
|  | *4 | 0,01851852 | 0,06666667 | -0,0481481 | 0,28913564 | 0,9655681 |
|  | *5 | 0,09259259 | 0,03333333 | 0,05925926 | 0,4138149 | 0,9655681 |
|  | *6 | 0,2962963 | 0,26666667 | 0,02962963 | 0,80712186 | 1 |
| CYP2C19 | *1 | 0,53703704 | 0,63333333 | -0,0962963 | 0,49160098 | 0,81933497 |
|  | *17 | 0,25925926 | 0,13333333 | 0,12592593 | 0,26756387 | 0,81933497 |
|  | *2 | 0,18518519 | 0,2 | -0,0148148 | 1 | 1 |
|  | *3 | 0 | 0,03333333 | -0,0333333 | 0,35714286 | 0,81933497 |
|  | *4 | 0,01851852 | 0 | 0,01851852 | 1 | 1 |
| CYP2C9 | *1 | 0,7962963 | 0,83333333 | -0,037037 | 0,77753019 | 1 |
|  | *11 | 0 | 0,03333333 | -0,0333333 | 0,35714286 | 1 |
|  | *2 | 0,14814815 | 0,13333333 | 0,01481481 | 1 | 1 |
|  | *3 | 0,05555556 | 0 | 0,05555556 | 0,54945216 | 1 |
| CYP2D6 | *1 | 0,25 | 0,23333333 | 0,01666667 | 1 | 1 |
|  | *10 | 0,05357143 | 0,1 | -0,0464286 | 0,41643589 | 1 |
|  | *2 | 0,28571429 | 0,3 | -0,0142857 | 1 | 1 |
|  | *3 | 0,01785714 | 0,03333333 | -0,0154762 | 1 | 1 |
|  | *34 | 0,01785714 | 0 | 0,01785714 | 1 | 1 |
|  | *4 | 0,16071429 | 0,2 | -0,0392857 | 0,76706711 | 1 |
|  | *41 | 0,10714286 | 0,06666667 | 0,04047619 | 0,70772054 | 1 |
|  | *5 | 0,03571429 | 0,03333333 | 0,00238095 | 1 | 1 |
|  | *6 | 0 | 0 | 0 | 1 | 1 |
|  | *9 | 0,03571429 | 0,03333333 | 0,00238095 | 1 | 1 |
|  | Dup1 | 0,01785714 | 0 | 0,01785714 | 1 | 1 |
|  | Dup2 | 0,01785714 | 0 | 0,01785714 | 1 | 1 |
| CYP3A4 | *1 | 0,88888889 | 0,9 | -0,0111111 | 1 | 1 |
|  | *1B | 0,05555556 | 0 | 0,05555556 | 0,54945216 | 0,99234158 |
|  | *22 | 0,05555556 | 0,1 | -0,0444444 | 0,66156105 | 0,99234158 |

Supplementary Table 10. Post-hoc allelic frequency comparisons between DEL and UPD populations. All comparisons were performed using Fisher exact test, with significance shown as p-values. False Discovery Rate Benjamini-Hochberg corrections were applied (q-values). Significant p- and q-values are shown in bold.

| **Gene** | **Phenotype** | **European Frequency** | **PWS Frequency** | **Delta Frequency** | **p-value** | **q-value (FDR-corrected)** |
| --- | --- | --- | --- | --- | --- | --- |
| CYP2B6 | UM | 0 | 0 | 0 | 1 | 1 |
|  | RM | 0,03703704 | 0 | 0,03703704 | 1 | 1 |
|  | NM | 0,48148148 | 0,4 | 0,08148148 | 0,74961656 | 1 |
|  | IM | 0,37037037 | 0,6 | -0,2296296 | 0,20230404 | 1 |
|  | PM | 0,11111111 | 0 | 0,11111111 | 0,54137631 | 1 |
|  | Other | 0 | 0 | 0 | 1 | 1 |
| CYP2C19 | UM | 0,03703704 | 0 | 0,03703704 | 1 | 1 |
|  | RM | 0,37037037 | 0,13333333 | 0,23703704 | 0,15797672 | 0,58038608 |
|  | NM | 0,25925926 | 0,46666667 | -0,2074074 | 0,19346203 | 0,58038608 |
|  | IM | 0,25925926 | 0,33333333 | -0,0740741 | 0,72595674 | 1 |
|  | PM | 0,07407407 | 0,06666667 | 0,00740741 | 1 | 1 |
|  | Other | 0 | 0 | 0 | 1 | 1 |
| CYP2C9 | NM | 0,59259259 | 0,66666667 | -0,0740741 | 0,74641957 | 1 |
|  | IM | 0,40740741 | 0,33333333 | 0,07407407 | 0,74641957 | 1 |
|  | PM | 0 | 0 | 0 | 1 | 1 |
|  | Other | 0 | 0 | 0 | 1 | 1 |
| CYP2D6 | UM | 0,07142857 | 0 | 0,07142857 | 0,53488372 | 1 |
|  | NM | 0,46428571 | 0,46666667 | -0,002381 | 1 | 1 |
|  | IM | 0,42857143 | 0,53333333 | -0,1047619 | 0,54014252 | 1 |
|  | PM | 0,03571429 | 0 | 0,03571429 | 1 | 1 |
|  | Other | 0 | 0 | 0 | 1 | 1 |
| CYP3A4 | NM | 0,77777778 | 0,8 | -0,0222222 | 1 | 1 |
|  | IM | 0,22222222 | 0,2 | 0,02222222 | 1 | 1 |

Supplementary Table 11. Post-hoc phenotypic frequency comparisons between DEL and UPD populations. All comparisons were performed using Fisher exact test, with significance shown as p-values. False Discovery Rate Benjamini-Hochberg corrections were applied (q-values). Significant p- and q-values are shown in bold.

| **Gene** | **Allele** | **European Frequency** | **PWS Frequency** | **Delta Frequency** | **p-value** | **q-value (FDR-corrected)** |
| --- | --- | --- | --- | --- | --- | --- |
| CYP2B6 | *1 | 0,55555556 | 0,5 | 0,05555556 | 0,66904965 | 0,78055792 |
|  | *19 | 0 | 0,02777778 | -0,0277778 | 0,4 | 0,78055792 |
|  | *2 | 0,03703704 | 0,08333333 | -0,0462963 | 0,38539236 | 0,78055792 |
|  | *22 | 0 | 0,02777778 | -0,0277778 | 0,4 | 0,78055792 |
|  | *4 | 0,01851852 | 0,05555556 | -0,037037 | 0,56149132 | 0,78055792 |
|  | *5 | 0,09259259 | 0,08333333 | 0,00925926 | 1 | 1 |
|  | *6 | 0,2962963 | 0,22222222 | 0,07407407 | 0,47591145 | 0,78055792 |
| CYP2C19 | *1 | 0,53703704 | 0,66666667 | -0,1296296 | 0,27624804 | 0,66666667 |
|  | *17 | 0,25925926 | 0,11111111 | 0,14814815 | 0,109623 | 0,54811499 |
|  | *2 | 0,18518519 | 0,19444444 | -0,0092593 | 1 | 1 |
|  | *3 | 0 | 0,02777778 | -0,0277778 | 0,4 | 0,66666667 |
|  | *4 | 0,01851852 | 0 | 0,01851852 | 1 | 1 |
| CYP2C9 | *1 | 0,7962963 | 0,77777778 | 0,01851852 | 1 | 1 |
|  | *11 | 0 | 0,02777778 | -0,0277778 | 0,4 | 1 |
|  | *2 | 0,14814815 | 0,13888889 | 0,00925926 | 1 | 1 |
|  | *3 | 0,05555556 | 0,05555556 | 0 | 1 | 1 |
| CYP2D6 | *1 | 0,25 | 0,21052632 | 0,03947368 | 0,80499442 | 1 |
|  | *10 | 0,05357143 | 0,13157895 | -0,0780075 | 0,26190772 | 1 |
|  | *2 | 0,28571429 | 0,26315789 | 0,02255639 | 1 | 1 |
|  | *3 | 0,01785714 | 0,02631579 | -0,0084586 | 1 | 1 |
|  | *34 | 0,01785714 | 0 | 0,01785714 | 1 | 1 |
|  | *4 | 0,16071429 | 0,23684211 | -0,0761278 | 0,42692761 | 1 |
|  | *41 | 0,10714286 | 0,07894737 | 0,02819549 | 0,73491986 | 1 |
|  | *5 | 0,03571429 | 0,02631579 | 0,0093985 | 1 | 1 |
|  | *6 | 0 | 0 | 0 | 1 | 1 |
|  | *9 | 0,03571429 | 0,02631579 | 0,0093985 | 1 | 1 |
|  | Dup1 | 0,01785714 | 0 | 0,01785714 | 1 | 1 |
|  | Dup2 | 0,01785714 | 0 | 0,01785714 | 1 | 1 |
| CYP3A4 | *1 | 0,88888889 | 0,89473684 | -0,005848 | 1 | 1 |
|  | *1B | 0,05555556 | 0 | 0,05555556 | 0,26469183 | 0,66252935 |
|  | *22 | 0,05555556 | 0,10526316 | -0,0497076 | 0,44168623 | 0,66252935 |

Supplementary Table 12. Post-hoc allelic frequency comparisons between DEL and Non-DEL populations. All comparisons were performed using Fisher exact test, with significance shown as p-values. False Discovery Rate Benjamini-Hochberg corrections were applied (q-values). Significant p- and q-values are shown in bold.

| **Gene** | **Phenotype** | **European Frequency** | **PWS Frequency** | **Delta Frequency** | **p-value** | **q-value (FDR-corrected)** |
| --- | --- | --- | --- | --- | --- | --- |
| CYP2B6 | UM | 0 | 0 | 0 | 1 | 1 |
|  | RM | 0,03703704 | 0 | 0,03703704 | 1 | 1 |
|  | NM | 0,48148148 | 0,44444444 | 0,03703704 | 1 | 1 |
|  | IM | 0,37037037 | 0,5 | -0,1296296 | 0,53914304 | 1 |
|  | PM | 0,11111111 | 0 | 0,11111111 | 0,26363636 | 1 |
|  | Other | 0 | 0,05555556 | -0,0555556 | 0,4 | 1 |
| CYP2C19 | UM | 0,03703704 | 0 | 0,03703704 | 1 | 1 |
|  | RM | 0,37037037 | 0,11111111 | 0,25925926 | 0,08585601 | 0,36564716 |
|  | NM | 0,25925926 | 0,5 | -0,2407407 | 0,12188239 | 0,36564716 |
|  | IM | 0,25925926 | 0,33333333 | -0,0740741 | 0,73945226 | 1 |
|  | PM | 0,07407407 | 0,05555556 | 0,01851852 | 1 | 1 |
|  | Other | 0 | 0 | 0 | 1 | 1 |
| CYP2C9 | NM | 0,59259259 | 0,55555556 | 0,03703704 | 1 | 1 |
|  | IM | 0,40740741 | 0,44444444 | -0,037037 | 1 | 1 |
|  | PM | 0 | 0 | 0 | 1 | 1 |
|  | Other | 0 | 0 | 0 | 1 | 1 |
| CYP2D6 | UM | 0,07142857 | 0 | 0,07142857 | 0,50786309 | 0,93545236 |
|  | NM | 0,46428571 | 0,36842105 | 0,09586466 | 0,56127142 | 0,93545236 |
|  | IM | 0,42857143 | 0,63157895 | -0,2030075 | 0,2375239 | 0,93545236 |
|  | PM | 0,03571429 | 0 | 0,03571429 | 1 | 1 |
|  | Other | 0 | 0 | 0 | 1 | 1 |
| CYP3A4 | NM | 0,77777778 | 0,78947368 | -0,0116959 | 1 | 1 |
|  | IM | 0,22222222 | 0,21052632 | 0,01169591 | 1 | 1 |

Supplementary Table 13. Post-hoc phenotypic frequency comparisons between DEL and Non-DEL populations. All comparisons were performed using Fisher exact test, with significance shown as p-values. False Discovery Rate Benjamini-Hochberg corrections were applied (q-values). Significant p- and q-values are shown in bold.
